# Common genetic and clinical risk factors: Association with fatal prostate cancer in the Cohort of Swedish Men

**DOI:** 10.1101/2020.09.16.20192666

**Authors:** Minh-Phuong Huynh-Le, Roshan Karunamuni, Chun Chieh Fan, Wesley K. Thompson, Kenneth Muir, Artitaya Lophatananon, Karen Tye, Alicja Wolk, Niclas Håkansson, Ian G. Mills, Ole A. Andreassen, Anders M. Dale, Tyler M. Seibert, The PRACTICAL Consortium

## Abstract

**Background:** Clinical variables—age, family history, genetics—are used for prostate cancer risk stratification. Recently, polygenic hazard scores (PHS46, PHS166) were validated as associated with age at prostate cancer diagnosis. While polygenic scores are associated with all prostate cancer (not specific for fatal cancers), PHS46 was also associated with age at prostate cancer death. We evaluated if adding PHS to clinical variables improves associations with prostate cancer death.

**Methods:** Genotype/phenotype data were obtained from a nested case-control Cohort of Swedish Men (n=3,279; 2,163 with prostate cancer, 278 prostate cancer deaths). PHS and clinical variables (family history, alcohol intake, smoking, heart disease, hypertension, diabetes, body mass index) were tested via univariable Cox proportional hazards models for association with age at prostate cancer death. Multivariable Cox models with/without PHS were compared with log-likelihood tests.

**Results:** Median age at last follow-up/prostate cancer death were 78.0 (IQR: 72.3-84.1) and 81.4 (75.4-86.3) years, respectively. On univariable analysis, PHS46 (HR 3.41 [95%CI 2.78-4.17]), family history (HR 1.72 [1.46-2.03]), alcohol (HR 1.74 [1.40-2.15]), diabetes (HR 0.53 [0.37-0.75]) were each associated with prostate cancer death. On multivariable analysis, PHS46 (HR 2.45 [1.99-2.97]), family history (HR 1.73 [1.48-2.03]), alcohol (HR 1.45 [1.19-1.76]), diabetes (HR 0.62 [0.42-0.90]) all remained associated with fatal disease. Including PHS46 or PHS166 improved multivariable models for fatal prostate cancer (*p*<10^-15^).

**Conclusions:** PHS had the most robust association with fatal prostate cancer in a multivariable model with common risk factors, including family history.

**Impact:** Adding PHS to clinical variables may improve prostate cancer risk stratification strategies.

## Introduction

Prostate cancer is a common malignancy that causes significant morbidity and mortality in men worldwide^1^. Risk-stratified screening has shown promise for identification of men most likely to develop prostate cancer^2–5^. Currently, estimates of a man’s risk may include clinical variables associated with prostate cancer, including family history, age, and race/ethnicity^6–11^. Other risk factors have also been reported^12,13^. Genetic factors may provide additional analytical and objective information^2,14,15^, possibly improving risk stratification beyond clinical variables alone.

A polygenic hazard score (PHS)—the weighted sum of approximately 50 single-nucleotide polymorphisms (SNPs)—has been developed and validated as independently associated with age at prostate cancer diagnosis. Though the association was not specific for fatal cancers, PHS was also associated with age at prostate cancer death^2,16,17^. The goal of the present work was to determine whether PHS improves stratification of risk for prostate cancer death beyond that achieved with clinical variables available in the Cohort of Swedish Men (COSM)^18^. COSM is a prospective, longitudinal, population-based study with participant-level demographic and clinical information. Data available include the development (or not) of prostate cancer and over two decades of mature follow-up data and death information^18^, affording an opportunity for comparison of genetic risk and clinical variables for associations with prostate cancer death.

## Methods

### The COSM Study

COSM is a longitudinal, population-based study designed to study associations of lifestyle with morbidity and mortality^18^. Men were surveyed at study onset in 1997, and in 2008 and 2009. Between 2005-2008, all the men still alive in the cohort were asked to give a saliva sample for genotyping through a self-administered postal collection^19^. A nested case-control study identified prostate cancer cases and controls. Prostate cancer cases included all the men with genotype data available and a diagnosis of prostate cancer made by the end of 2011. Men without prostate cancer at last follow-up were selected as controls, matched to the age distribution of the cases. There were 3,279 men (2,163 prostate cancer cases, 1,116 controls) in this COSM dataset. Prior work suggested this dataset would have adequate power for the present analyses^20^.

We identified the following clinical variables available in the COSM cohort: family history of prostate cancer (yes or no), alcohol intake (yes or no), smoking status (never smoker, current daily smoker, and ex-smoker), history of heart disease (yes or no), history of hypertension (yes or no), history of diabetes (clinically diagnosed by doctor; yes or no), and body mass index (BMI; kg/m^2^). Family history was taken as presence or absence of a first degree relative with a prostate cancer diagnosis. The number of relatives has also been associated with prostate cancer risk^6,10,21^, but there were not enough available data in the COSM dataset to include this variable in the present analysis.

### Polygenic Hazard Score (PHS)

A version of the original PHS with 46 SNPs^2,16^ (PHS46) was previously shown to be independently associated with any, aggressive, and fatal prostate cancers in men of European, African, and Asian genetic ancestry in a heterogeneous, multi-ethnic dataset, which included the COSM data analyzed here. The combination of PHS46 and family history in multivariable models performed better than family history alone for all endpoints^16^.

Here, we tested the performance of the adapted PHS for association with age at prostate cancer death in the COSM dataset, specifically, taking advantage of the rich clinical information, long follow-up, and population-based design of the COSM study. PHS46 was calculated for all patients in COSM and used as the sole covariate in Cox proportional hazards regressions for age at prostate cancer death. Secondarily, we also evaluated PHS46 association with age at any and aggressive prostate cancer diagnosis. As in prior reports^2,16^, aggressive prostate cancer was defined as disease typically requiring treatment: Gleason score ≥7, clinical stage T3-T4, PSA concentration ≥10ng/mL, or nodal/distant metastasis. Separate Cox proportional hazards regressions were performed for age at diagnosis of any or aggressive prostate cancer. All *p*-values reported were truncated at <10^-16 2^. As Cox proportional hazards results may be biased by a higher number of cases in the dataset compared to the general population, sample weight corrections were applied to all Cox proportional hazards models in this study using Swedish population data, as described previously^2,16,22^.

Recognizing that more SNPs associated with prostate cancer have been reported since the development of the original and adapted PHS^23^, a PHS model has also recently been developed that incorporates 166 total SNPs (PHS166)^24^, drawing from the SNPs in PHS46 and SNPs reported in a meta-analysis of genome-wide association studies^23^. PHS166 improved the hazard ratios (HRs) for prostate cancer over PHS46. The COSM data were included in the SNP selection and model weight estimation of PHS166, so there may be some within-sample overestimation of performance. With that caveat, we additionally evaluated PHS166 here for association with fatal prostate cancer in COSM in models accounting for clinical variables.

### Clinical variables and PHS – association with fatal prostate cancer

Univariable Cox proportional hazards models were used to determine whether individual clinical variables were independently associated with age at prostate cancer death. If a participant did not have the clinical variable of interest available, they were not included in those specific Cox models. Those clinical variables with *p*<0.05 for univariable prediction of age of diagnosis of any prostate cancer were combined in a multivariable model. To evaluate whether addition of genetic risk improved prediction over clinical variables alone, a log-likelihood test compared the multivariable model with and without PHS46 as a predictor. Finally, the multivariable model with PHS46 was compared to the univariable model with PHS46 as the sole predictor of age of diagnosis of any prostate cancer, again by log-likelihood test. Significance for comparison of multivariable models was set at α=0.05.

The above multivariable analyses were repeated, with PHS166 used instead of PHS46.

## Results

### COSM participants

**Table 1** describes the cohort. There were 278 deaths from prostate cancer, as well as 1,403 cases meeting clinical criteria for aggressive disease. The median age at last follow-up was 78 years (IQR: 72.3, 84.1), and the median age at prostate cancer death was 81.4 years (IQR: 75.4, 86.3).

**Table 1.** Participant characteristics, n=3,279.

| **<u>Participants</u>** | **<u>Number</u>** |
| --- | --- |
| Controls | 1,116 |
| Prostate cancer cases | 2,163 |
| Aggressive prostate cancer cases <sup>a</sup> | 1,403 |
| Fatal prostate cancer cases | 278 |
| **<u>Age Demographics</u>** | **<u>Median (IQR) <sup>b</sup></u>** |
| Age at prostate cancer diagnosis | 72.4 (66.2, 78.5) |
| Age at last follow up | 78.0 (72.3, 84.1) |
| Age at prostate cancer death | 81.4 (75.4, 86.3) |
| **<u>Polygenic Hazard Scores</u>** | **<u>Median (IQR)</u>** |
| PHS46 overall | 0.4863 (0.2976, 0.6949) |
| PHS46 in fatal prostate cancer cases | 0.5035 (0.3306, 0.7051) |
| PHS46 in non-fatal prostate cancer cases | 0.4850 (0.2968, 0.6935) |
| PHS166 overall | 0.0148 (-0.2306, 0.2508) |
| PHS166 in fatal prostate cancer cases | 0.0770 (-0.1548, 0.2957) |
| PHS166 in non-fatal prostate cancer cases | 0.0042 (-0.2380, 0.2472) |
| **<u>Participants with Known Clinical Variables Information</u>** | **<u>Number</u>** |
| Family history | 2,453 |
| Heart disease | 3,279 |
| Hypertension | 3,279 |
| Alcohol intake | 3,268 |
| Smoking | 3,237 |
| Diabetes | 3,279 |
| Body mass index | 3,132 |

<sup>a</sup> Aggressive prostate cancer defined as Gleason score $\geq 7$ , PSA $\geq 10$ ng/mL, T3-T4 stage, nodal metastases, or distant metastases.
<sup>b</sup> IQR: interquartile range

### PHS in COSM

PHS46 was associated with age at prostate cancer death in the COSM dataset (HR 3.41 [95% CI 2.78, 4.17], *z*=12, *p*<10^-16^). PHS46 was also associated with any (HR 5.35 [4.97, 5.76], *z*=45, *p*<10^-16^) and aggressive (HR 3.78 [3.45, 4.13], *z*=29, *p*<10^-16^) prostate cancer, consistent with prior findings that SNP associations are not specific for aggressive forms of cancer.

PHS166 was also associated with age at prostate cancer death (HR 7.15 [95% CI 5.95, 8.58], *z*=21, *p*<10^-15^).

### Clinical variables and PHS – Association with fatal prostate cancer

On univariable analysis, family history of prostate cancer (HR 1.72 [95% CI 1.46, 2.03], *z*=6.5, *p*<10^-9^), alcohol intake (HR 1.74 [1.40, 2.15], *z*=5.1, *p*<10^-6^), and diabetes history (HR 0.53 [0.37, 0.75], *z*=-3.6, *p*=0.003) were each associated with prostate cancer death; these were included in the multivariable models. Smoking history (HR 0.96 [0.84, 1.10], *z*=-0.6, *p*=0.56 for ex-smokers; HR 0.92 [0.79, 1.08], *z*=-1.0, *p*=0.34 for current daily smokers), heart disease (HR 0. 97 [0.83, 1.13], *z*=-0.4, *p*=0.68), hypertension (HR 1.06 [0.93, 1.21], *z*=0.9, *p*=0.30), and BMI (HR 0.99 [0.97, 1.01], *z*=-0.8, *p*=0.45) were not associated with prostate cancer death.

In the multivariable model with all clinical variables (with univariable significance), each variable remained independently associated with prostate cancer death. Family history of prostate cancer (HR 1.70 [1.44, 2.01], *z*=6.3, p<10^-9^) and history of alcohol intake (HR 1.44 [1.17, 1.78], *z*=3.5, *p*=0.0005) were associated with increased risk of prostate cancer death. Diabetes history (HR 0.63 [0.43, 0.92], *z*=-2.4, *p*=0.014) was associated with decreased risk of prostate cancer death.

Adding PHS46 improved multivariable model performance. PHS46, family history, alcohol intake, and diabetes history each remained independently significant in the multivariable model, with PHS (HR 2.45 [1.99, 2.97], *z*=8.6, *p*<10^-15^) and family history (HR 1.73 [1.48, 2.03], *z*=6.5, *p*<10^-10^) having the strongest associations with prostate cancer death (**Table 2**). A log-likelihood test confirmed model improvement with inclusion of genetic risk, compared to only clinical variables (*p*<10^-15^). Similarly, the full multivariable model was significantly better than PHS alone (log-likelihood test, *p*<10^-10^).

**Table 2.** Multivariable Cox proportional hazards model with both the polygenic hazard score (PHS46) and all clinical variables with significant univariable association with prostate cancer death.

| **Clinical variable** | **$\beta$** | **HR (95% CI)** | **z-score** | ***p*-value** |
| --- | --- | --- | --- | --- |
| PHS46 | 0.90 | 2.45 (1.99, 2.97) | 8.7 | $<10^{-15}$ |
| Family history of prostate cancer | 0.55 | 1.73 (1.48, 2.03) | 6.5 | $<10^{-10}$ |
| Alcohol intake | 0.37 | 1.45 (1.19, 1.76) | 3.5 | 0.0004 |
| Diabetes | -0.48 | 0.62 (0.42, 0.90) | -2.5 | 0.012 |

Multivariable models with PHS166 were also improved with addition of PHS166. PHS166, family history, and alcohol intake each remained independently significant in the multivariable model, with PHS166 (HR 5.48 [4.54, 6.61], *z*=17.9, *p*<10^-15^) having the strongest association with prostate cancer death (**Table 3**). Diabetes history was not independently significant in the multivariable model using PHS166. A log-likelihood test confirmed model improvement with inclusion of genetic risk, compared to only clinical variables (p<10^-15^). Similarly, the full multivariable model was significantly better than PHS alone (log-likelihood test, *p*<10^-7^).

**Table 3.** Multivariable Cox proportional hazards model with PHS166 and all clinical variables with significant univariable association with prostate cancer death.

| **Clinical variable** | **$\beta$** | **HR (95% CI)** | **z-score** | ***p*-value** |
| --- | --- | --- | --- | --- |
| PHS166 | 1.70 | 5.48 (4.54, 6.61) | 17.9 | $<10^{-15}$ |
| Family history of prostate cancer | 0.39 | 1.48 (1.25, 1.75) | 4.5 | $<10^{-5}$ |
| Alcohol intake | 0.42 | 1.52 (1.22, 1.88) | 3.9 | $<10^{-4}$ |
| Diabetes | -0.31 | 0.73 (0.49, 1.07) | -1.6 | 0.11 |

## Discussion

In a nested case-control study from a large, population-based, longitudinal Swedish cohort^18^, we found that a multivariable model accounting for both clinical and genetic factors best stratified men for risk of fatal prostate cancer. Specifically, the addition of PHS significantly improved associations for age at prostate cancer death over that of the clinical variables alone. PHS46 had the most robust effect in the multivariable model, after controlling for other clinical variables in the COSM dataset. Incorporating more SNPs, PHS166 improved on the performance of PHS46 and yielded a similar pattern of genetics as an important risk factor for fatal prostate cancer. Our work suggests that a multifactorial approach combining genetics and medical history might be useful for prostate cancer risk stratification.

In the present analysis, multiple clinical variables were associated with fatal prostate cancer. We found that family history of prostate cancer, alcohol intake, and a lack of diabetes were associated with increased likelihood of prostate cancer death. These results are consistent with previous investigations into prostate cancer risk factors. Family history is a well-known risk factor for prostate cancer already used in clinical decision-making^6,10,11,21^. Alcohol consumption has been linked with prostate cancer risk in a recent meta-analysis^12^. Diabetes history, on the other hand, may actually reduce a man’s risk of prostate cancer^25–27^. While each of these epidemiological findings are consistent with the present findings, this is the first study to specifically assess for association with age at prostate cancer death^12,25,28^. Age is a critical consideration in prostate cancer decision-making^29^, especially as medical co-morbidities and competing risk of death increase with aging^30^. At the same time, prostate cancer incidence rises exponentially with age and is typically more aggressive when diagnosed at an older age^31,32^.

Among the variables evaluated in this study, family history of prostate cancer was strongly and independently associated with prostate cancer death, consistent with prior reports and with the current use of family history in clinical risk stratification^6,7,10,11,28,33^. In fact, on multivariable analysis with PHS46, family history yielded the second highest hazard ratio (1.73) for prostate cancer death, second only to that of PHS46 (2.45). Family history does not appear to be an adequate surrogate for an individual’s genetic risk, however, as PHS46 was also independently associated with prostate cancer death, indicating PHS and family history offer complementary risk information.

The present work is limited to clinical variables available in the COSM dataset. Other prostate cancer risk factors have been reported previously, including the number of affected family members in those men with a family history of prostate cancer^6,10,21^. Additionally, this is a study of a single population; while the PHS has been validated in a multi-ethnic dataset for association with fatal prostate cancer^16^, the relationships reported here between clinical variables and PHS may not generalize to other populations and warrant further study^34^. The focus of this study was the important endpoint of prostate cancer death, but, consistent with prior work, germline SNP associations here are not specific to fatal or aggressive prostate cancer. Finally, the performance of PHS166 may have been influenced by incorporation of COSM data in the prior development of the PHS166 model (done via a LASSO-regularized Cox regression framework)^24^; however, potential bias is likely low. Despite these limitations, our results indicate that polygenic scores like PHS may significantly improve strategies to identify men at highest (or lowest) risk of dying from prostate cancer.

## Conclusions

PHS and clinical variables afford complementary information regarding risk of fatal prostate cancer. In a multivariable model, PHS, family history, alcohol intake, and history of diabetes were each independently associated with prostate cancer death. PHS had the largest hazard ratio for fatal prostate cancer among these variables. Combining genetic and clinical factors should be studied as a strategy to improve risk-stratification for prostate cancer screening.

## Data Availability

Most of the data used here are available on the dbGaP platform managed by the National Institutes of Health (https://www.ncbi.nlm.nih.gov/gap, Study Accession number phs001391.v1.p1). We obtained our dataset for this work from the Prostate Cancer Association Group to Investigate Cancer Associated Alterations in the Genome (PRACTICAL) consortium, which consists of a collaborative group of researchers, each of whom retains ownership of their contributed data. Members of the consortium can use pooled data via proposals that are reviewed by the Data Access Committee. Approved proposals are then sent to the principal investigators of the PRACTICAL member studies, each of whom may opt to participate or not in the specific request. Readers interested in participating in the PRACTICAL consortium and gaining access to member data may find information about application at: http://practical.icr.ac.uk.

## Funding

This study was funded in part by a grant from the United States National Institute of Health/National Institute of Biomedical Imaging and Bioengineering (#K08EB026503), United States Department of Defense (#W81XWH-13-1-0391), the Research Council of Norway (#223273), KG Jebsen Stiftelsen, and South East Norway Health Authority. The content is solely the responsibility of the authors and does not necessarily represent the official views of any of the funding agencies, who had no role in the design and conduct of the study; collection, management, analysis, and interpretation of the data; preparation, review, or approval of the manuscript; and decision to submit the manuscript for publication.

## Conflicts of Interest

TM Seibert reports honoraria from Multimodal Imaging Services Corporation for imaging segmentation and honoraria from Varian Medical Systems and WebMD, Inc. for educational content. OA Andreassen has a patent application # U.S. 20150356243 pending; AM Dale also applied for this patent application and assigned it to UC San Diego. AM Dale has additional disclosures outside the present work: founder, equity holder, and advisory board member for CorTechs Labs, Inc.; advisory board member of Human Longevity, Inc.; recipient of nonfinancial research support from General Electric Healthcare.

## Data & Code Availability

Most of the data used here are available on the dbGaP platform managed by the National Institutes of Health (https://www.ncbi.nlm.nih.gov/gap, Study Accession number phs001391.v1 .p1). We obtained our dataset for this work from the Prostate Cancer Association Group to Investigate Cancer Associated Alterations in the Genome (PRACTICAL) consortium, which consists of a collaborative group of researchers, each of whom retains ownership of their contributed data. Members of the consortium can use pooled data via proposals that are reviewed by the Data Access Committee. Approved proposals are then sent to the principal investigators of the PRACTICAL member studies, each of whom may opt to participate or not in the specific request. Readers interested in participating in the PRACTICAL consortium and gaining access to member data may find information about application at: http://practical.icr.ac.uk. Code used for this work are stored in an institutional repository and will be shared upon request to the corresponding author.

## Notes

### Author Declarations

University of California San Diego

